# Tracking a decade of structural changes in preclinical and prodromal Alzheimer’s disease: insights from amyloid-β pathology

**DOI:** 10.1101/2025.09.25.25336640

**Authors:** Ting Qiu, Zhen-Qi Liu, Jonathan Gallego-Rudolf, Manon Edde, Alex Valcourt Caron, Yuanchao Zhang, M. Mallar Chakravarty, Jean-Paul Soucy, D. Louis Collins, Judes Poirier, John Breitner, Gemma Salvadó, Alexa Pichet Binette, Maxime Descoteaux, Sylvia Villeneuve, the PREVENT-AD Research Group

## Abstract

Alzheimer’s disease (AD) pathology is typically associated with reduced brain volume and cortical thickness, interpreted as neurodegeneration. However, several cross-sectional studies in cognitively unimpaired individuals have paradoxically reported larger brain volume and thicker cortex in the presence of amyloid-β (Aβ), suggesting that Aβ–brain structure associations may not follow a simple linear pattern early in the disease. We leveraged over a decade of longitudinal data from the PREVENT-AD cohort (N=367, mean follow-up 7.17 years, range 0–11.27 years) to investigate how Aβ burden related to both cross-sectional levels and longitudinal changes in brain volume, cortical thickness, and cortical tissue mean diffusivity (MD_T_) derived from free-water corrected diffusion tensor imaging. We examined associations separately in individuals below (Aβ−) and above (Aβ+) the Aβ-positivity threshold, tested for nonlinearity across all participants, and aligned structural trajectories to the estimated years from Aβ-positivity onset. We found that higher Aβ was associated with larger brain volume, higher cortical thickness and lower MD_T_ in regions such as the fusiform gyrus, supramarginal gyrus, hippocampal volume, inferior parietal and middle temporal cortex in the Aβ− group. The opposite associations were found in the Aβ+ group. Across all participants, volume and thickness showed an inverse U-shaped relationship with Aβ, while MD_T_ followed a U-shaped pattern. Longitudinally, the rate of cortical thickness change showed an inverse U-shaped association with Aβ, whereas Aβ burden was associated with faster volume loss and greater MD_T_ increase. When structural measures were aligned to the estimated time of Aβ positivity onset, volume and thickness increased years before the expected Aβ positivity onset and declined thereafter, while MD_T_ showed no association with time relative to Aβ positivity onset. Our results help reconcile inconsistencies across prior studies and suggest that brain structural changes related to Aβ pathology start years before Aβ positivity onset and follow nonlinear trajectories. These early structural changes might be due to pathological processes such as neuroinflammatory swelling early in the course of the disease.

## 1 INTRODUCTION

Alzheimer’s Disease (AD) involves a long preclinical stage during which amyloid-β (Aβ) plaques and neurofibrillary tau tangles accumulate in the brain, ultimately leading to neurodegeneration and cognitive decline.^1–4^ Numerous studies have reported that Aβ and tau pathology are associated structural brain deterioration, including reductions in cortical thickness and hippocampal volume which can be measured *in vivo* using magnetic resonance imaging (MRI)^5,6^. However, emerging evidence suggests that changes in brain structure during preclinical AD may not follow a monotonic decline.^7–11^ Instead, they may exhibit biphasic trajectories that could challenge the identification of early AD-related brain changes and bias the monitoring and interpretation of preventive interventions. Reliable indicators of early brain changes in AD need to be identified to guide timely and effective interventions, before irreversible neuronal damage occurs.^12–15^

While most studies have reported reduced brain volume and cortical thickness in individuals expressing AD pathology, some cross-sectional studies conducted in cognitively unimpaired individuals have observed the opposite: larger gray matter volume and increased cortical thickness being associated with greater Aβ pathology.^7,8^ This findings suggest a biphasic trajectory, where structural measures first change in one direction and later reverse as pathology advances.^7,16,17^ Such biphasic patterns have also been shown in individuals with autosomal dominant AD (ADAD) using cortical mean diffusivity (MD), a diffusion MRI diffusion tensor imaging (DTI) biomarker sensitive to microscopic tissue changes in the grey matter. One of the first interpretation for these unexpected results was that people with a larger premorbid brain size can tolerate more pathology before cognitive function is affected, also conceptualized as higher “brain reserve”.^18^ An alternative hypothesis is that pathological processes such as Aβ accumulation, swelling and neuroinflammation create an expansion of the grey matter, resulting in greater volume and cortical thickness in individuals who do not yet experience neurodegeneration.^19,20^ Together, these findings suggest a potential non-linear relationship between AD pathology and structural changes, with early transient alterations preceding a shift toward neurodegeneration as pathology progresses.

We analyzed over 10 years of longitudinal neuroimaging data from the Pre-symptomatic Evaluation of Experimental or Novel Treatments for AD (PREVENT-AD) cohort to investigate the association between Aβ pathology and brain structure, focusing on grey matter volume, cortical thickness, and cortical tissue mean diffusivity (MD_T_, where T stands for tissue) derived from free-water corrected DTI. These structural measures capture complementary aspects of brain integrity: volume reflects macroscopic changes, while cortical thickness and cortical MD_T_ are both sensitive to microstructural tissue integrity.^8,21,22^ We tested whether a biphasic association exists between Aβ burden and structural brain trajectories in the early phases of AD. First, we applied partial least square analysis (PLS) separately in individuals with and without significant levels of Aβ, referred to as Aβ negative (Aβ−) and Aβ positive (Aβ+) groups. We used partial least squares (PLS) analyses applied to cross-sectional and longitudinal structural measures to identify brain regions of interest (ROIs) showing opposite associations with Aβ pathology. We then extracted Aβ levels and structural measures from the PLS-derived ROIs and tested for non-linear associations across all participants. Lastly, to map individual trajectories relative to disease progression, we estimated each participant’s Aβ positivity onset age and computed years relative to Aβ positivity,^23^ in order to test whether structural markers reflect brain expansion/reduction as a function of estimated years from Aβ positivity.

## 2 METHODS

### 2.1 Participants

We used data from PREVENT-AD dataset, a monocentric longitudinal study initiated in 2011 that comprises data from 387 cognitively unimpaired older adults with a familial history of AD dementia.^24,25^ The specific inclusion and exclusion criteria for the PREVENT-AD study are detailed in Tremblay-Mercier et al.^24^

We focused on a subsample of 241 participants who underwent both Aβ PET imaging and structural/diffusion MRI. For the cross-sectional analysis, we selected the MRI visit acquired closest to the PET (average time between PET and MRI: 0.17 ± 1.27 years). While continuous Aβ values were used in most of analyses, for the PLS we divided participants into low (Aβ−) and high (Aβ+) Aβ deposition groups using a predefined global Aβ threshold based on early Aβ accumulating regions.^26,27^ The PET scans and the related MRI were not necessarily performed at the beginning of the study and therefore 35 participants had already developed MCI by the time they did the PET or MRI scan. Among these, we excluded 17 MCI participants who were Aβ−, as their cognitive impairment was unlikely due to AD and may reflect other diseases. The final cross-sectional sample was comprised of 224 participants: 136 cognitively unimpaired Aβ− individuals and 88 Aβ+ individuals (70 cognitively unimpaired and 18 MCI; **Table 1**; **Figure S1**). Among the 224 participants included in the cross-sectional analyses, 214 had at least one follow-up MRI scan (mean follow-up time of 7.66 years ± 2.04; range 0.99–11.20 years; mean number of scans = 4.26 ± 1.45; range = 2–7), and 105 had a follow-up Aβ PET scan (mean time: 4.36 ± 0.57 years; range 1.59–6.29 years; **Figure S1**).

**Table 1.**
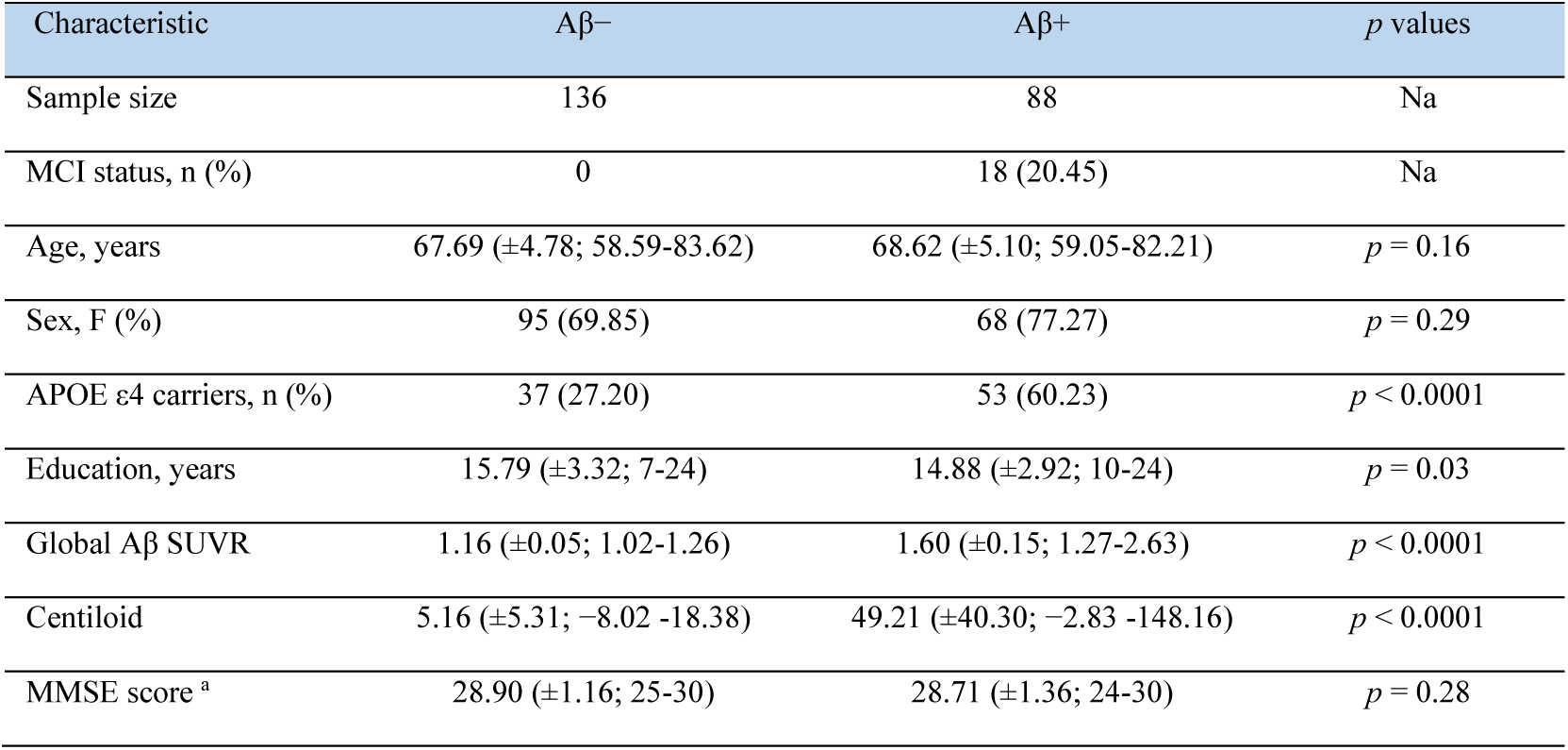
Demographic, pathological, and clinical characteristics of the study. . Data is depicted as the mean (standard deviation; range), except in the case of categorical variables, for which the count and percentage are presented. Abbreviations: MCI, mild cognitive impairment; F, female; APOE, apolipoprotein E; MMSE, Mini-Mental State Examination. ^a^ MMSE score was available for 219 participants.

All participants provided informed consent for their de-identified data to be used for research purposes. Our research protocol is compliant with the Declaration of Helsinki and was reviewed and approved by the Institutional Review Board at McGill University (Protocol #A05-B16-11B and #A05-M55-11B).

### 2.2 Image acquisition

All MRI data (N=995) were acquired at the Douglas Mental Health University (Montreal, Canada). In November 2018, the scanner was upgrade from a Siemens 3T Tim Trio to a Siemens 3T Prisma, dividing the dataset into two acquisition stages: 56% of the scans were acquired before the upgrade (Stage1: 2011-2017), and 44% after (Stage2: 2019-2023; **Figure S2**). In both stages, T1-weighted (T1w) MRI scans were obtained using a three-dimensional (3D) MPRAGE sequence. Diffusion-weighted MRI (DWI) were collected using a Pulse Gradient Spin Echo (PGSE) EPI sequence, with single-shell acquisitions before the upgrade and multi-shell acquisitions after. Detailed acquisition parameters for T1w and DWI scans collected from both stages are provided in **Supplementary Table 1**. PET scans (N = 329; 224 baseline and 105 follow-up) were performed on a brain-dedicated Siemens/CTI high-resolution research tomograph (HRRT; scanner resolution: 2.5 mm) at the McConnell Brain Imaging Centre of the Montreal Neurological Institute (Montreal, Canada). Aβ PET scans were acquired 40-70 minutes after injection of [^18^F]NAV4694 (220 ± 22 MBq). Image acquisition comprised six 5-minute frames acquired in this time window, along with an attenuation scan. Images were then reconstructed using a 3D ordinary Poisson ordered subset expectation maximum (OP-OSEM) algorithm with 10 iterations and 16 subsets.^28,29^

### 2.3 MRI processing

T1w MRI data were processed using FreeSurfer version 5.3.0 (http://surfer.nmr.mgh.harvard.edu/). The FreeSurfer pipeline consists of automated steps to reconstruct the grey/white interface and pial surface of the cortex. Cortical thickness maps were obtained by measuring the distance between the grey/white boundary and pial surfaces^30,31^ The cortical surface was segmented into 68 ROIs as defined by the Desikan-Killiany (DK) atlas, and cortical volume and thickness were extracted for each ROI. Hippocampal volumes were obtained from the subcortical segmentation output of FreeSurfer. Regional volumes were divided by total intracranial volume (TIV), a commonly used approach to account for interindividual differences in head size.^32^

DWI data were processed using Tractoflow 2.4.3. Detailed information about the pipeline can be found at https://github.com/scilus/tractoflow.^33,34^ Briefly, the preprocessing steps involved denoising, correction for field inhomogeneity, geometric distortions, eddy-current effects, brain extraction, normalization, and up sampling to 1 mm.^28,33^ Our measure of interest, MD_T_, was computed using the freely available Freewater_flow pipeline (https://github.com/scilus/freewater_flow). This method fits a regularized bi-tensor model in each voxel to quantify the isotropic diffusion of the free water (FW) fraction and tissue compartment (see our previous work for more details^28^). Once the MD_T_ maps were computed, they were warped to the native space of the T1w scans acquired during the same session. To reduce partial volume effects,^8^ the warped MD_T_ maps were projected into individual’s cortical surface using the FreeSurfer *mri_vol2surf* command. We then extracted the cortical MD_T_ values for each of the 68 cortical regions defined by the DK atlas for subsequent analyses.

Since 56% of the MRI data were acquired before the scanner upgrade and the remaining data were collected after the upgrade, brain volume, cortical thickness and MD_T_ for each region were harmonized across scanners using the longitudinal ComBat method.^35^ Prior work has shown that scanner upgrade from Siemens Tim Trio to Prisma may affect MRI-derived measures,^36^ so harmonization was applied to limit potential scanner-related variability in longitudinal data while preserving biological variability.^35^ As an example, brain volume, cortical thickness and MD_T_ values within a representative region were plotted before and after harmonization (**Figure S3**).

### 2.4 PET processing

All PET images were processed using our in-house pipeline (https://github.com/villeneuvelab/vlpp). Briefly, the Aβ-PET images were realigned, averaged, co-registered to their closest available T1w MRI scans and segmented using the DK atlas in Freesurfer.^30^ The registered PET images were masked to exclude CSF activity and smoothed using a 6 mm Gaussian kernel. Standardized uptake value ratios (SUVRs) were calculated for each region by normalizing the signal intensity of that region to the cerebellar cortex.^37^ Global Aβ burden was calculated as the weighted average SUVR across a set of bilateral medial and lateral frontal, parietal, and temporal regions.^27^ Participants with a global Aβ uptake greater than our previously defined threshold of 1.26 were classified as Aβ+.^26^

Longitudinal Aβ accumulation trajectories were estimated using a sampled iterative local approximation algorithm (SILA), which is publicly available (https://github.com/Betthauser-Neuro-Lab/SILA-AD-Biomarker). This nonparametric modeling approach consists of three main steps: (1) sampling the rate of Aβ accumulation at discrete age intervals based on available longitudinal PET data; (2) applying a smoothing technique to reduce noise and to refine the estimated accumulation rates over time; and (3) performing numerical integration to generate a nonparametric curve that models Aβ accumulation as a function of age. More details regarding this method can be found in Betthauser et al.^23^ Using the fitted Aβ curve, each participant’s estimated age at Aβ positivity onset was determined by identifying the age at which the modeled Aβ trajectory crossed the SUVR threshold of 1.26. The number of years from estimated Aβ+ (years**_→_** _Aβ+_) was then computed by subtracting the estimated age of Aβ+ onset from the participant’s age at each MRI timepoint. This allowed us to estimate the timing of Aβ positivity onset and further model structural changes relative to the inferred course of Aβ pathology.

### 2.5 Statistical analyses

We evaluated differences in demographic, pathological, and clinical measurements between the Aβ− and Aβ+ groups using independent sample t-tests for continuous variables and Chi-square tests for categorical variables.

#### 2.5.1 Cross-sectional analyses

##### 2.5.1.1 Multivariate PLS analysis

To examine group-specific multivariate associations between Aβ pathology and brain structure, we performed PLS analyses using the *pyls* Python package (https://github.com/netneurolab/pypyls), fitted separately for individuals with low (Aβ−) and high (Aβ+) Aβ burden. This group wise approach was based on the premise that structural changes associated with Aβ accumulation may differ depending on whether individuals are at an early or later stages of pathology. Given that PLS captures the dominant pattern of covariance within a dataset,^38^ pooling together all participants could mask unique associations where contrasting effects exist between Aβ− and Aβ+ individuals. For each group, we performed three separate PLS analyses to assess the associations between regional Aβ SUVR values and (1) brain volume, (2) cortical thickness and (3) cortical MD_T_. Regional Aβ SUVR values derived from the DK atlas served as predictor variables, while the corresponding regional measures of volume, thickness and MD_T_ were used as outcomes in each PLS model (**Figure 1A**).

**Figure 1.**
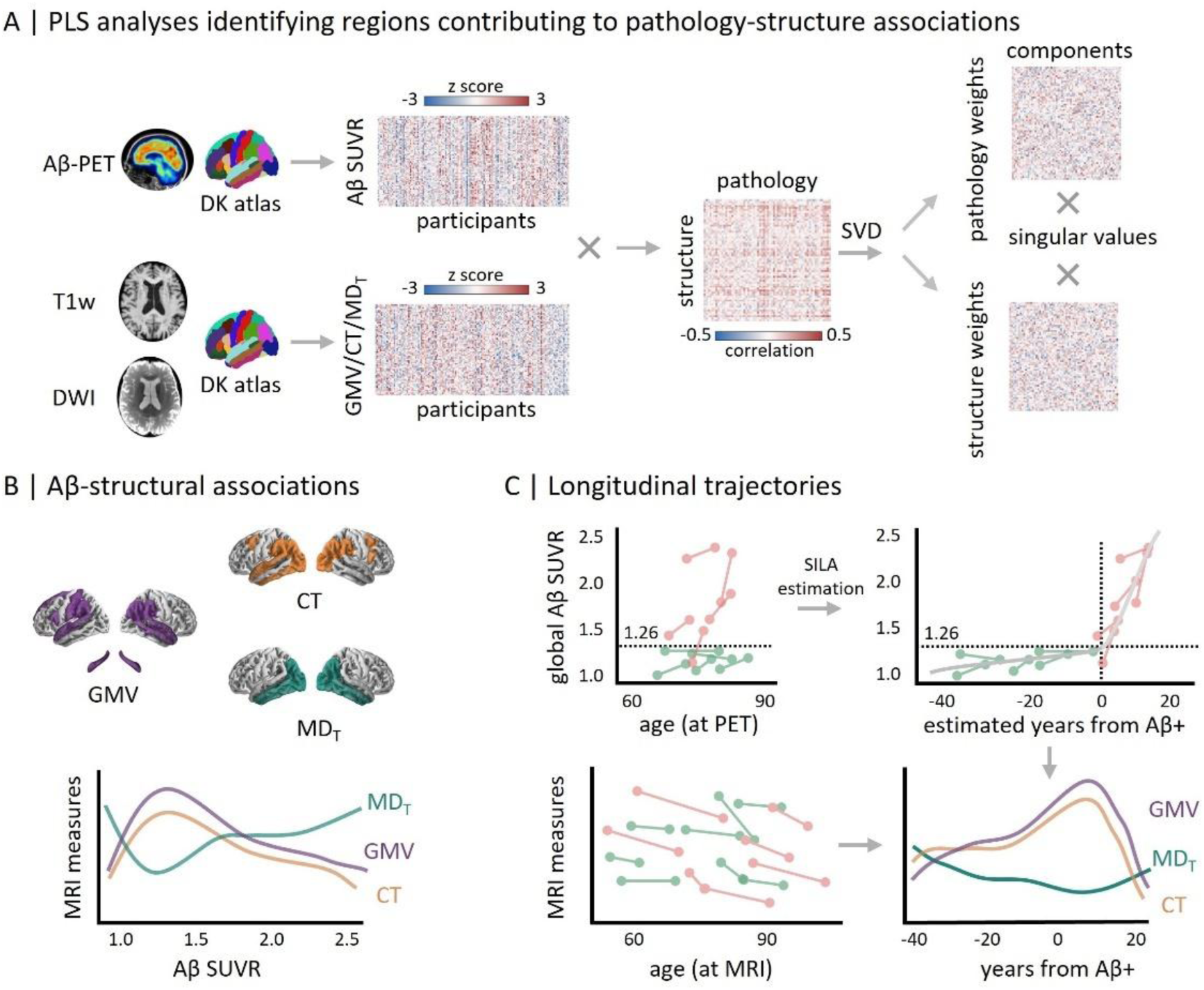
Methodology Overview. **(A)** PLS analyses were performed to examine the strongest multivariate patterns of covariance between Aβ deposition and structural measures (GMV/CT/MD_T_), and between Aβ deposition and slope of structural measures in Aβ− and Aβ+ groups. **(B)** GAM models were fitted to examine the association between Aβ burden and structural measures or structural changes (slope) within key regions identified from PLS analyses. All participants were analyzed as a single group. **(C)** Longitudinal Aβ-PET data were used to train the SILA algorithm to estimate the years from Aβ positivity; the estimated age of Aβ positivity onset for each participant was calculated by subtracting the estimated time from Aβ positivity from their chronological age at the reference scan; years of Aβ positivity at longitudinal MRI scans were calculated by subtracting the age of Aβ positivity onset age from the age at each MRI; GAMM models were fitted to model the longitudinal structural changes with the time from Aβ positivity. DWI, diffusion weighted MRI; DK, Desikan–Killiany; GMV, grey matter volume; CT, cortical thickness; MD_T_, free-water-corrected mean diffusivity; HV, hippocampal volume; SVD, singular value decomposition; LVs, latent variables; GAM, generalized additive model; PLS, partial least squares; SILA, sampled iterative local approximation; GAMM, generalized additive mixed model.

To assess the significance and robustness of the identified associations, we evaluated each latent variable (LV) derived from each PLS using permutation tests (10,000 iterations), retaining only those with permutation-based *p* value < 0.05. Each LV represents a distinct pattern of covariance between Aβ pathology and brain structure. The first LV captures the strongest associations, while subsequent LVs account for additional, statistically independent patterns.^38,39^ These patterns are mathematically orthogonal, which implies that each LV captures different aspects of the Aβ–structure relationship.^38,39^ For each LV, pathological and structural scores were computed by projecting the original data onto their corresponding weight vectors. These scores represent how strongly each participant expresses the pattern of the pathology–structure associations.^38,39^ Correlating these two scores across participants reflects the strength of the multivariate relationship between Aβ pathology and brain structure captured by the PLS model.

We assessed the stability of regional contributions to each LV using bootstrap resampling (10,000 iterations). For each regional measure, a bootstrap ratio (BSR) was computed as the ratio of the original PLS weight divided by its bootstrap-estimated standard error.^38,39^ To further examine the spatial expression of the latent patterns, we computed pathological loadings (correlations between pathological features and PLS-derived structural scores) and structural loadings (correlations between structural features and PLS-derived pathological scores). These loadings reflect the strength of the association between each regional feature (e.g., Aβ-SUVR, brain volume or thickness, MD_T_) and the underlying latent pattern.^38,39^ The 95% confidence intervals (CIs) for the loadings were estimated from the bootstrap distribution. Features with a BSR ≥ 1.96 (equivalent to bootstrap-estimated 95% CIs excluding zero) were considered stable contributors to the covariance pattern, that is, features with a strong and reliable contribution to the multivariate pattern. Key ROIs were defined as the regions where the same modality-specific regional measures met BSR (i.e., stability) criteria in both Aβ− and Aβ+ PLS analyses. Subsequent analyses focused on these overlapping ROIs.

##### 2.5.1.2 Non-linear associations between Aβ pathology and structural measures

While the PLS analyses allowed us to identify ROIs showing opposite Aβ–structural relationships when stratifying participants by Aβ-PET status, we also wanted to test the complexity (or non-linearity) of these associations at a whole group level. To do so, we first extracted the average Aβ SUVR, brain volume, cortical thickness, and cortical MD_T_ values from the corresponding PLS-derived ROIs (**Figure 1B**). Bilateral hippocampal volumes were also extracted as a stand-alone measure, considering its early involvement in AD and its clinical relevance for staging disease severity.^40,41^ Lastly, we extracted brain volume, cortical thickness, and cortical MD_T_ values from a sensorimotor regions ROI to be used as a control region for which we expected linear or at least less complex association with Aβ, given that Aβ deposition occurs in this region only late in the disease.^42^

To assess the non-linearity of the associations between Aβ pathology and structural measures within the PLS-derived ROIs, we used generalized additive models (GAMs; *mgcv* R package v1.8-41) to flexibly model potential non-linear relationships (**Figure 1B**). Each GAM included age and sex as covariates:

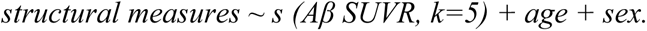

Here, *s (.)* denotes a penalized smoothing spline with a basic dimension *k=5*. Smoothing parameters were optimized using restricted maximum likelihood (REML). Effective degrees of freedom (EDF) were reported to quantify model non-linearity, with EDF >1 indicating deviation from a linear relationship and with higher values showing greater complexity.

In addition to modeling Aβ SUVR as a continuous predictor, we also fit separate models using rank-transformed Aβ levels to assess whether observed non-linearity was consistent when minimizing the influence of outliers and skewed distributions.

#### 2.5.2 Longitudinal analyses

##### 2.5.2.1 Multivariate PLS analysis using longitudinal structural measures

To examine group-specific associations between Aβ pathology and longitudinal changes in brain structure, we repeated our PLS analyses described above, replacing the cross-sectional measures with individual slopes (rate of change) of regional brain volume, cortical thickness, and cortical MD_T_. The longitudinal slope for each measure was calculated using separate linear mixed effects models (LMMs; *lme4* R package v1.1-34), with random intercepts and slopes included to account for within-subject variability over time. For each group (Aβ− and Aβ+), three separate PLS models related regional Aβ SUVR values to the longitudinal slopes of regional brain volume, cortical thickness, and cortical MD_T_. Significant latent variables were identified via permutation testing (10,000 iterations), and stable regional contributors via bootstrap ratios (|BSR| ≥ 1.96). Key longitudinal slope PLS-derived ROIs were defined as regions meeting stability criteria in both groups for the same modality.

##### 2.5.2.2 Non-linear associations between Aβ pathology and longitudinal changes in structural measures

We then examined potential non-linear associations between Aβ burden and longitudinal structural changes across the whole cohort. For each participant, longitudinal slopes of brain volume, cortical thickness, and cortical MD_T_ values were extracted within the longitudinal PLS-derived ROIs and sensorimotor control regions, as well as slopes of bilateral hippocampal volume. These slopes were then modeled as a function of Aβ SUVR using GAMs, with age and sex included as covariates:

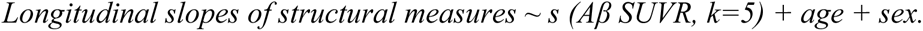

Analogous to the cross-sectional analyses, we repeated the models using rank-transformed Aβ levels as the predictor to assess robustness to outliers and skewed distributions.

#### 2.5.3 Longitudinal structural trajectories aligned to estimated years from Aβ positivity

One limitation of human studies is that enrolled participants are not necessary at the same disease stage and/or they do not all have the same level of pathology.^43^ To model structural trajectories along the course of Aβ progression, we used SILA-estimated time from Aβ positivity (*years****_→_*** *_Aβ+_*) as a temporal proxy for cumulative Aβ burden (**Figure 1C**). The SILA framework infers individualized Aβ accumulation timelines from longitudinal Aβ-PET data in a subset of participants (see SILA estimation in **section 2.4**). Longitudinal structural changes were modeled using generalized additive mixed models (GAMMs; *gamm4* R package v0.2-6). Each model included a smooth term for SILA-estimated *years****_→_*** *_Aβ+_,* and adjusted for age and sex as covariates:

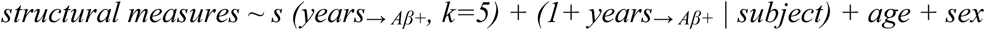

The smooth terms s (.) used penalized regression splines with a basis dimension of k = 5. Subject-specific random intercepts and slopes *(1+ years****_→_*** *_Aβ+_ | subject)* were included in the model to account for individual variability in baseline structural measures and rates of change. Smoothing parameters were optimized via the REML method.

We modeled longitudinal changes in brain structure within combined ROIs, defined as the union of the regions identified by cross-sectional and longitudinal PLS analyses. As sensitivity analyses, we repeated all models using each ROI set separately and additionally applied them to sensorimotor regions for comparison. We first fit the GAMM within Aβ− and Aβ+ groups to examine stage-dependant differences in structural trajectories, then applied the same model to the full sample to characterize the overall trajectory patterns.

All statistical analyses were conducted using R software (v4.2.1), except for the PLS analyses, which were performed in Python (v3.8.12). Statistical significance was determined using two-sided *p*-values ≤ 0.05, adjusted for multiple comparisons using false discovery rate (FDR).

## 3 RESULTS

### 3.1 Demographic characteristics of the groups

Participants’ demographics are summarized in **Table 1**. Among the 224 participants with both Aβ-PET and structural/diffusion MRI data, 136 participants were classified as Aβ− and 88 as Aβ+. The mean age (Aβ−: 67.69±4.78 years; Aβ+: 68.62±5.10 years; *p* = 0.16) and proportions of female participants (77.27% vs. 69.85%; *p* = 0.29) were comparable between the two groups. The Aβ+ group showed higher prevalence of APOE ε4 carriers (60.23% vs. 27.20%; *p* < 0.0001) and lower educational attainment (14.88±2.92 vs. 15.79±3.32 years; *p* = 0.03).

### 3.2 Cross-sectional analyses

#### 3.2.1 Multivariate PLS

In the Aβ− group, two significant LVs were identified for each of the three PLS models: Aβ–volume (LV1: 60.27% cross-block covariance, permutation *p* < 0.0001; LV2: 9.49% covariance, *p* = 0.003), Aβ–cortical thickness (LV1: 59.29% cross-block covariance, *p* = 0.006; LV2: 12.69% covariance, *p* = 0.03), and Aβ–MD_T_ models (LV1: 81.49% covariance, *p* < 0.0001; LV2: 9.09%, *p* = 0.03). We focused on LV1 in each model, as it accounted for the largest proportion of variance; LV2 results are reported in **Figure S9.** In the Aβ–volume model, higher Aβ levels in the precuneus, supramarginal, inferior parietal, and superior parietal cortices were associated with higher grey matter volume of the precuneus, supramarginal, inferior parietal, hippocampus, and medial temporal regions. A similar spatial pattern was observed in the Aβ–cortical thickness model, where higher Aβ deposition in regions typically affected early in AD such as the lateral and medial orbitofrontal, precuneus, superior parietal, and inferior temporal cortices, was associated with higher cortical thickness in the supramarginal, entorhinal, fusiform, and superior frontal cortices. In contrast, the Aβ–MD_T_ model showed that higher Aβ levels were linked to lower MD_T_, especially in posterior and prefrontal regions such as the precuneus, inferior temporal, superior frontal, and entorhinal cortices (**Figure 2A**). We also observed significant correlations between weighted scores of pathological and structural features for the Aβ–volume (*r* = 0.37; *p* < 0.0001), the Aβ–cortical thickness (*r* = 0.31; *p* < 0.001), and the Aβ–MD_T_ (*r* = 0.36; *p* < 0.0001) models.

**Figure 2.**
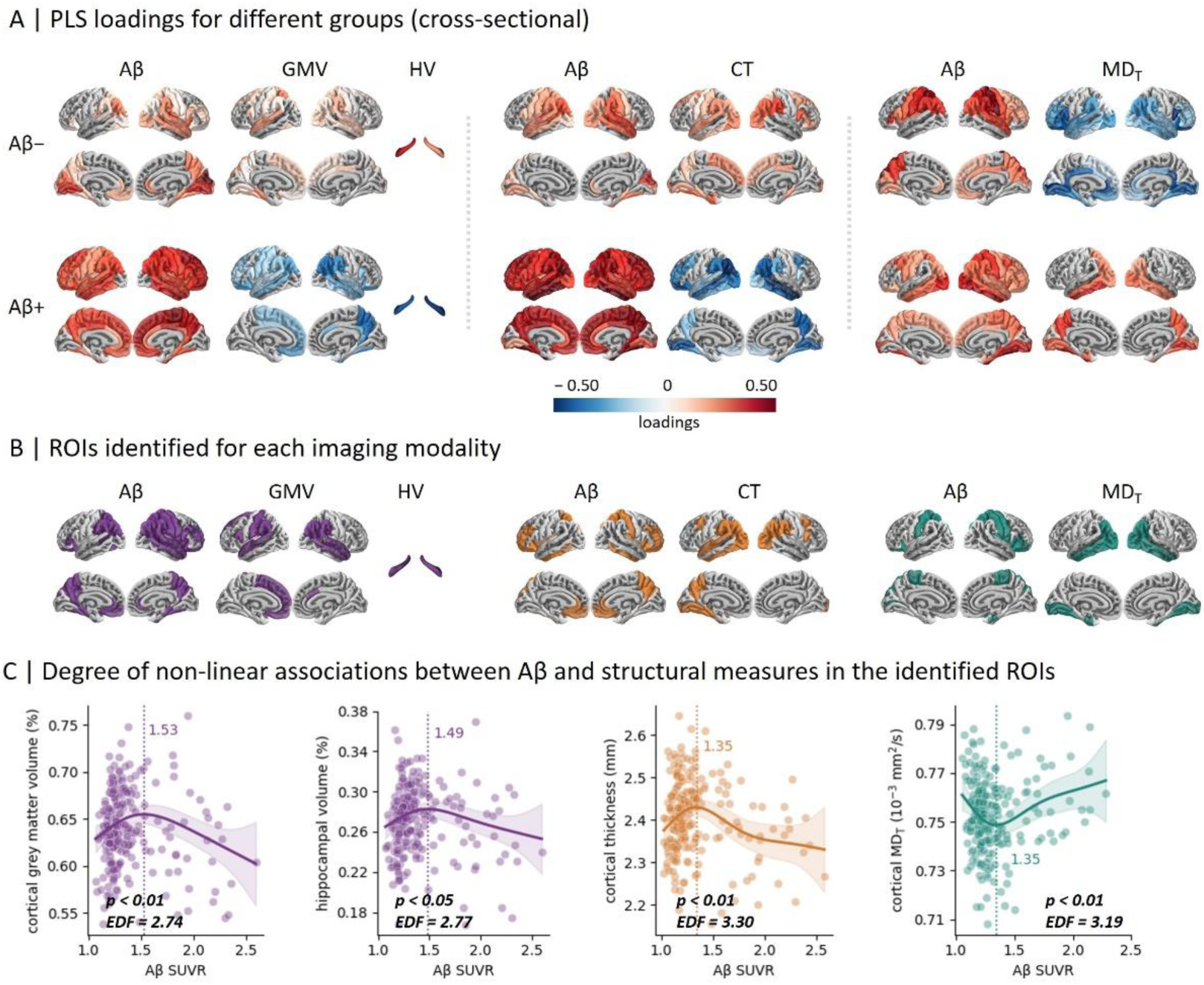
PLS loadings across groups and associations between AD pathology and structural measures within the cross-sectional PLS-derived regions. **(A)** PLS loadings from the first latent variable are displayed on brain surface maps for regional Aβ and structural measures (brain volume, cortical thickness and MD_T_). Loadings are color-coded to indicate the direction and reliability of their contributions: positive (red), negative(blue), or null (gray). Regions with loadings of the same sign indicate positive covariance, whereas regions with opposite signs indicate negative covariance. **(B)** Regions that contributed reliably in Aβ− and Aβ+ groups are shown for each imaging modality and referred to as overlapping ROIs. **(C)** Degree of non-linear associations between Aβ and structural measures were analyzed using GAMs, adjusting for age and sex; EDF > 1 indicates deviation from linearity, with larger EDF values reflecting greater complexity in the association. The top row shows results for continuous Aβ SUVR, while the bottom row displays associations using rank-transformed Aβ levels. The vertical dotted line marks the Aβ level at which the structural measure peaks. GMV, grey matter volume; HV, hippocampal volume; CT, cortical thickness; MD_T_, free-water-corrected mean diffusivity; GAM, generalized additive model; PLS, partial least squares; EDF, estimated degrees of freedom.

In the Aβ+ group, a single LV emerged in each model: Aβ–volume (LV1: 91.04% covariance, *p* = 0.002), Aβ–cortical thickness (LV1: 92.47% covariance, *p* = 0.002), and Aβ–MD_T_ (LV1: 89.36% covariance, *p* = 0.02). Higher global Aβ levels were linked to lower gray matter volume, particularly in the lateral and medial temporal cortices (e.g., fusiform, inferior temporal, parahippocampal, hippocampus), as well as the inferior parietal and superior frontal regions. The Aβ–cortical thickness model showed a similar pattern of cortical thinning, largely overlapping with regions implicated in the Aβ–volume model. In the Aβ–MD_T_ model, higher global Aβ levels were associated with higher MD_T_ in the fusiform, precuneus, entorhinal, and inferior temporal regions. Compared to the Aβ− group, structural alterations in the Aβ+ group affected a broader set of regions in volume and thickness, while MD_T_ changes were more restricted (**Figure 2A**). We found significant correlations between weighted scores of pathological and structural features for the Aβ– volume (*r* = 0.36; *p* < 0.001), the Aβ–cortical thickness (*r* = 0.39; *p* < 0.001), and the Aβ– MD_T_ (*r* = 0.28, *p* < 0.01) models.

**Figure 2A** shows the PLS loadings across cortical regions, reflecting the contribution of each region to the latent pattern. The corresponding BSR maps, showing the stability of these contributions, are provided in **Figure S5**. Loadings and BSR were highly correlated, showing strong consistency in terms of strength and stability (**Figure S6**). For each measure, we identified key ROIs as regions that met the stability criterion (|BSR| ≥ 1.96) in both the Aβ− and Aβ+ groups. These ROIs are summarized in **Figure 2B**.

#### 3.2.2 Non-linear associations within PLS-derived ROIs

In this set of ROIs derived from the cross-sectional PLS analyses, we observed that grey matter volume followed an inverse U-shaped association with Aβ burden (EDF = 2.74, *p* = 0.006, R^2^_adj_ = 0.25). At lower Aβ levels, greater burden was linked to larger volumes, peaking at SUVR ≈ 1.53, whereas at higher levels it was related to lower volumes. A similar pattern was found for the hippocampal volume (EDF = 2.77, *p* = 0.03, R^2^_adj_ = 0.29), with a peak at Aβ SUVR ≈ 1.49. Cortical thickness in the PLS-derived ROIs also followed an inverse U-shaped association (EDF = 3.30, *p* = 0.004, R^2^_adj_ = 0.18), with the highest value observed around SUVR ≈ 1.35. In contrast, cortical MD_T_ showed a U-shaped relationship with Aβ burden (EDF = 3.19, *p* = 0.005, R^2^_adj_ = 0.24). At lower Aβ levels, greater Aβ burden was associated with lower cortical MD_T_, reaching a minimum near SUVR ≈ 1.35. At higher Aβ levels, greater Aβ burden was associated with higher MD_T_. Similar results were obtained when using rank-transformed Aβ levels instead of SUVRs, supporting the robustness of these associations (**Figure S7**). In contrast, no significant associations were found in the sensorimotor control regions (**Figure S8**).

### 3.3 Longitudinal analyses

#### 3.3.1 Multivariate PLS analysis using structural longitudinal slopes

In both the Aβ− and Aβ+ groups, only the first LV was significant across all PLS models when incorporating longitudinal structural measures.

In the Aβ− group, LV1 explained 63.75% of the cross-block covariance for the Aβ–volume slope model (*p* < 0.0001), 50.20% for the Aβ–cortical thickness slope model (*p* < 0.0001), and 60.02% in the Aβ–MD_T_ slope model (*p* < 0.0001). Higher Aβ burden was associated with faster volume loss in the precuneus, supramarginal, inferior parietal, and medial temporal cortices, except in the isthmus of the cingulate gyrus, where higher Aβ was linked to slower decline. For cortical thickness, higher Aβ burden was associated with slower thinning in the fusiform, supramarginal, and parahippocampal regions. Higher Aβ was also related to greater increases in MD_T_ in the posterior and temporal cortices. Significant correlations between weighted scores of pathological and structural features were observed for the longitudinal Aβ–volume (*r* = 0.33; *p* < 0.0005), Aβ–cortical thickness (*r* = 0.38; *p* < 0.0001), and Aβ–MD_T_ (*r* = 0.41; *p* < 0.0001) models.

In the Aβ+ group, LV1 accounted for 90.52% of the cross-block covariance in the Aβ– volume longitudinal model (*p* < 0.0001), 92.44% in the Aβ–cortical thickness longitudinal model (*p* < 0.0001), and 81.03% in the Aβ–MD_T_ longitudinal model (*p* < 0.0001). Higher Aβ burden was associated with faster decline for both brain volume and cortical thickness, with the most prominent effects observed in the lateral and medial temporal, parietal, and frontal cortices. Greater Aβ burden was associated with greater increases in MD_T_ longitudinal slopes in the interior and middle temporal regions, while slower increases in MD_T_ in cingulate regions (**Figure 3A**). Significant correlations between the weighted pathology and structural scores were observed in the longitudinal Aβ–volume (*r* = 0.43; *p* < 0.0001), the Aβ–cortical thickness (*r* = 0.43; *p* < 0.0001), and the Aβ–MD_T_ (*r* = 0.49; *p* < 0.0001) models.

**Figure 3.**
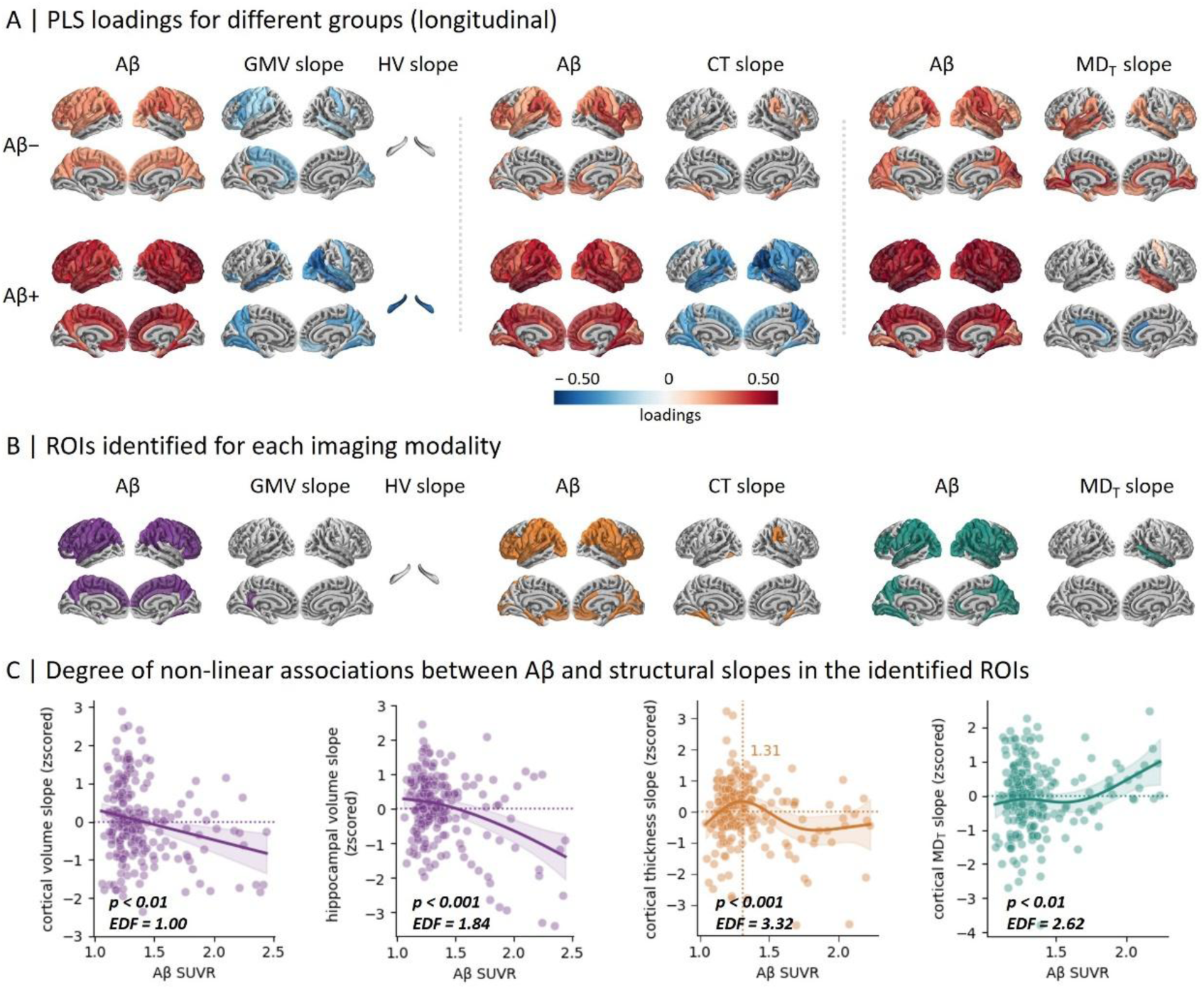
PLS loadings across groups and associations between Aβ pathology and slopes of structural measures within the slope PLS-derived regions. **(A)** PLS loadings from the first latent variable are displayed on brain surface maps for regional Aβ and slopes of structural measure (brain volume, cortical thickness, and MD_T_). (B) Regions that contributed reliably in both Aβ− and Aβ+ groups are shown for each imaging modality and referred to as slope-based ROIs. (C) Degree of non-linear associations between Aβ burden and the slopes of structural measures were analyzed using GAMs, adjusting for age and sex; EDF > 1 indicates deviation from linearity, with larger EDF values reflecting greater complexity in the association. The top row shows results for continuous Aβ SUVR, while the bottom row displays associations using rank-transformed Aβ levels. The vertical dotted line marks the Aβ level at which the slope of the structural measure peaks, and a horizontal line denotes zero slope (no change). GMV, grey matter volume; HV, hippocampal volume; CT, cortical thickness; MD_T_, free-water-corrected mean diffusivity; GAM, generalized additive model; PLS, partial least squares; EDF, estimated degrees of freedom.

**Figure 3A** shows the longitudinal PLS loadings across cortical regions, reflecting their contribution to the latent pattern. Key ROIs were defined as regions meeting the stability criterion (|BSR| ≥ 1.96) in both the Aβ− and Aβ+ groups and are summarized in **Figure 3B**.

#### 3.3.2 Non-linear associations within longitudinal PLS-derived ROIs

Within these ROIs, longitudinal changes in cortical grey matter volume (EDF = 1.00, p = 0.001, R^2^_adj_ = 0.05) and hippocampal volume (EDF = 1.84, p < 0.001, R^2^_adj_ = 0.20) both showed a monotonic decrease in relation to Aβ burden. Longitudinal changes in cortical thickness exhibited a non-linear association with Aβ burden (EDF = 3.32, *p* < 0.001, R^2^_adj_ = 0.19). At lower Aβ levels, cortical thickness longitudinal slopes increased and became positive, peaking at SUVR ≈ 1.32, whereas at higher Aβ levels, the slopes became negative. MD_T_ showed increasing rate of change with greater Aβ burden (EDF = 2.62, *p* < 0.01, R^2^_adj_ = 0.26). These patterns remained consistent when using rank-transformed Aβ levels, as shown in (**Figure S7**). No significant associations were observed in the sensorimotor control regions (**Figure S8**).

### 3.4 Longitudinal structural trajectories align with estimated years from Aβ positivity

We modeled structural trajectories as a function of SILA-estimated years since Aβ positivity onset (*years**_→_** _Aβ+_*), a temporal proxy reflecting Aβ burden severity, with negative values denoting years preceding Aβ positivity onset and positive values reflecting post-onset progression (**Figure 4A**; **Figure 4B**).

**Figure 4.**
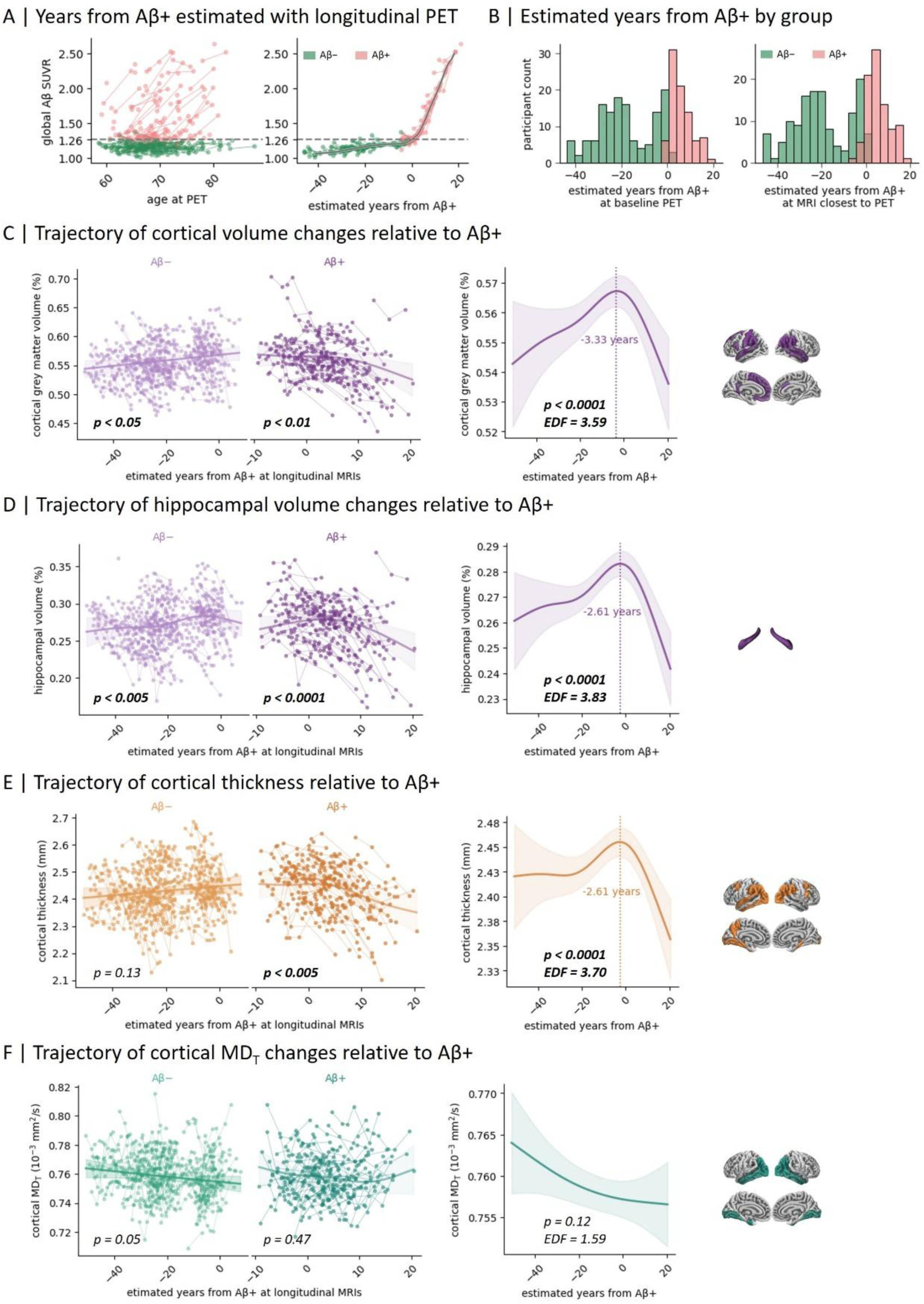
Longitudinal structural trajectories over time from Aβ positivity. **(A)** Longitudinal Aβ-PET data were used to estimate individual time from Aβ positivity onset using SILA algorithm. Left: longitudinal Aβ-PET SUVR plotted against age at Aβ PET scan. Right: SILA-estimated time from Aβ positivity plotted against observed Aβ SUVR. **(B)** Distributions of SILA-estimated time from Aβ positivity are shown for each group at baseline PET (left) and at the MRI scan closest to baseline PET (right). **(C-F)** Longitudinal trajectories of cortical grey matter volume, hippocampal volume, cortical thickness and MD_T_ within corresponding ROIs plotted as function of SILA-estimated years from Aβ positivity. Left panels: analyses stratified by Aβ− and Aβ+ groups. Right panels: trajectories with all participants combined. The vertical dotted line indicates the transition from increases to declines in volume and thickness. SILA, sampled iterative local approximation; MD_T_, free-water-corrected mean diffusivity.

We first examined longitudinal trajectories of cortical grey matter volume, hippocampal volume, cortical thickness and MD_T_ within the ROIs for individuals classified as low (Aβ−) and high (Aβ+) Aβ deposition groups. Cortical grey matter volume increased as time approached Aβ positivity in the Aβ− group (EDF = 1.00, p = 0.02, R²adj = 0.35) and decreased with increasing *years****_→_*** *_Aβ+_* in the Aβ+ group (EDF = 2.97, p = 0.009, R²adj = 0.22; **Figure 4 C**). Hippocampal volume showed the same pattern, increasing in the Aβ− group and declining with *years****_→_*** *_Aβ+_* in the Aβ+ group (**Figure 4 D**). Cortical thickness decreased over time in the Aβ+ group (EDF = 3.19, p < 0.001, R²adj = 0.19) but showed no correlation with *years****_→_*** *_Aβ+_* in the Aβ− group (**Figure 4 E)**. Cortical MD_T_ was not associated with *years****_→_*** *_Aβ+_* in either group (**Figure 4 F)**.

When modeling all participants together, cortical grey matter volume (EDF = 3.59, *p* = 0.0002, R^2^ _adj_ = 0.29), hippocampal volume (EDF = 3.83, *p* < 0.0001, R^2^ _adj_ = 0.31) and cortical thickness (EDF = 3.70, *p* < 0.0001, R^2^ _adj_ = 0.15) showed inverted-U trajectories with respect to *years****_→_*** *_Aβ+_*. These measures increased in the years leading up to Aβ positivity, peaking at approximately −3.33 years for cortical volume, −2.61 years for both hippocampal volume and cortical thickness, and declined thereafter. Again, cortical MD_T_ was not associated with *years****_→_*** *_Aβ+_* (**Figure 4**). Comparable non-linear patterns were observed when trajectories were modeled using cross-sectional and longitudinal PLS-derived ROIs separately (**Figures S9)**. Structural measures in the sensorimotor control regions were not associated with *years****_→_*** *_Aβ+_* (**Figures S10**).

## 4 DISCUSSION

Understanding how AD pathology drives brain structural changes over time is central to staging disease progression and identifying early intervention windows.^4,20,44^ Here we leveraged over 10 years of longitudinal MRI together with PET imaging to investigate how Aβ burden relates to changes in brain structure across the early phases of the AD continuum. Rather than relying on predefined brain regions,^5,23^ we used a data-driven approach (i.e., PLS) to identify spatial patterns of pathological–structural associations within individuals with low or subthreshold (Aβ−) versus high (Aβ+) Aβ burden, with the assumption that Aβ starts to accumulate in the subthreshold phase.^45,46^ Given the clinical importance of hippocampal volume in staging disease severity,^40,41^ we also assessed its association with Aβ burden separately. Within these ROIs, we found contrasting patterns between Aβ− vs Aβ+ individuals, with both brain volume and cortical thickness showing inverse U-shaped relationships with Aβ burden, while MD_T_ followed a U-shaped pattern. These curves reflect the same biphasic process, given that lower volume/thickness typically corresponds to higher diffusivity. Similar non-linear associations with Aβ pathology were also observed for hippocampal volumes and longitudinal changes in cortical thickness. To further characterize structural changes along the temporal course of Aβ accumulation, we estimated years to Aβ positivity using the SILA method and aligned structural measures as a function of this timeline. We found that brain volume and cortical thickness increased prior to Aβ positivity, peaked around the estimated point of positivity conversion, and then declined over the following decades. Together, these findings suggest that structural brain changes follow a dynamic biphasic trajectory, with distinct phases of alteration across early AD. They further support the emerging view that subthreshold Aβ levels are meaningful, as structural changes emerge before individuals reach the threshold for Aβ positivity.^46^ Our multivariate cross-sectional PLS analyses revealed stage-dependent spatial patterns of Aβ-related structural changes across our different imaging markers. Among Aβ− individuals, Aβ deposition was associated with higher brain volume and cortical thickness in precuneus, supramarginal, inferior parietal, and medial temporal regions, as well as lower MD_T_ in prefrontal and posterior areas, all regions known to be vulnerable to early Aβ accumulation.^27,47^ In contrast, Aβ+ individuals with higher Aβ burden showed lower volume and cortical thickness in parietal and medial temporal regions, along with higher MD_T_. While each marker showed distinct spatial profiles, the inferior parietal and medial temporal cortices were consistently involved across volume, thickness and MD_T_, suggesting that these regions may be particularly vulnerable to early Aβ-related processes in preclinical AD.^10,48^

The association between Aβ pathology and brain structure is stage-dependent, which may help reconcile inconsistencies across prior cross-sectional studies. Previous work have reported paradoxical findings when examining the relationship between AD pathology and structural measures,^22,49–53^ with some reporting cortical thinning or higher conventional MD in relation to AD pathology among MCI and/or AD patients,^50–52^ and others suggesting larger volume/thickness and lower MD in cognitively unimpaired participants with abnormal Aβ pathology.^11,17,53–55^ In our cross-sectional analyses using PLS-derived ROIs, we observed biphasic patterns: lower Aβ burdens correlated with both higher brain volume and cortical thickness and lower FW-corrected MD (MD_T_), whereas higher Aβ levels corresponded to lower volume and thickness and higher MD_T_. These findings are also consistent with previous studies conducted in patients with ADAD.^8,16^

Importantly, analyses of longitudinal structural measures also revealed stage-dependent association with Aβ burden, especially for changes in cortical thickness, which showed an inverse U-shaped association with Aβ burden. This biphasic pattern in cortical thickness change aligns with previous longitudinal work showing slower atrophy rates in early preclinical stages and accelerated atrophy in medial temporal regions at later stages, particularly once abnormal CSF Aβ and tau are both present.^16^ Additionally, a recent study found that cortical thickness differences emerge at least seven years before PET-detectable positivity: individuals who later become Aβ+ already show thicker cortex on MRI measures and slower thinning than Aβ− individuals,^56^ which is complementary with our findings.

Finaly, we applied the SILA algorithm to estimate individual-specific years to Aβ positivity (years→ Aβ+), enabling the temporal alignment of longitudinal MRI measures with inferred Aβ burden across disease progression. This approach provides a continuous proxy for Aβ burden severity,^23^ and allows structural trajectories to be modeled across the early phases of the disease. Using this framework, we modeled the longitudinal trajectories of brain volume, cortical thickness and MD_T_ within the PLS-derived ROIs, as a function of continuous estimates of Aβ progression (i.e., *years****_→_*** *_Aβ+_*). We found that brain volume and cortical thickness followed similar biphasic trajectories: both increased during the years prior to Aβ positivity onset, peaked around the estimated transition point, and declined thereafter. These results further support the notion that initial increases and subsequent decreases in brain volume/thickness signal different phases of the pathological process.^16^

These findings suggest a critical window around the onset of Aβ positivity, during which brain structure shifts from transient alterations to irreversible degeneration. Early interventions targeting this phase may therefore help prevent the acceleration of widespread neurodegeneration.^13,14,57^ Our study highlights the importance of modeling the temporal dynamics of longitudinal structural trajectories to better understand disease progression and optimize therapeutic strategies.^4^

The biphasic trajectories observed across structural markers likely reflect distinct neurobiological responses to Aβ pathology. In the early phase, Aβ accumulation might trigger a neuroinflammation process (and/or vice versa), as well as glial activation or transient neuronal hypertrophy, leading to increased volume/thickness and reduced MD_T_ in vulnerable regions.^12,17,53,58,59^ As Aβ burden progresses, chronic neuroinflammation and synaptic dysfunction may precipitate a breakdown of structural connections, resulting in progressive cortical thinning and elevated MD_T_.^58^ This interpretation is supported by evidence showing that early glial reactivity is likely initially associated with transient structural ‘pseudo-normalization’ via inflammatory process, before transitioning to irreversible neurodegeneration with advancing Aβ pathology.^60,61^ Comparable paradoxical increases in local brain volume have also been observed in AD mouse models, particularly in early life and under metabolic stressors such as high-fat diet,^62,63^ supporting the possibility that such expansions may represent early pathological processes rather than brain reserve.^18^

Another potential explanation for these paradoxical increases in volume/thickness is that the physical space occupied by Aβ plaques drives the changes observed in measures of brain structure. Post-mortem studies have shown that Aβ plaques can occupy up to 6% of cortical volume.^64–67^ Similarly, anti-amyloid trials have reported cerebral volume loss in treatment group after plaque removal, which has been interpreted as Aβ-removal-related pseudoatrophy.^67–69^ If removing Aβ can lead to reduced volume due to clearance of plaques and associated tissue responses, it is possible to hypothesize that Aβ accumulation overtime may contribute to subtle increases in volume/thickness during the early stages of the disease.^67^ This explanation is likely limited to early phases, as later decreases observed over time are more consistent with neurodegeneration. Other interpretations include the influence of vascular alterations, the distinct vulnerability of different cell types, and the inter-individual variability in resilience factors, which should be considered and investigated in future studies.^70–72^

This study has several strengths. By defining regions with stage-dependent pathological– structural associations based on a data-driven approach, we identified key ROIs where biphasic trajectories are most pronounced, highlighting the most prominent structural changes in the early stages of the AD continuum. The availability of over 10 years of longitudinal MRI data allows us to characterize structural changes across an extended preclinical and prodromal window. To address the fact that PET was not available for every MRI scan, we applied SILA algorithm to estimate time from Aβ positivity onset, which allowed us to align longitudinal MRI changes with inferred Aβ progression. Despite these strengths, the PET system has lower effective resolution, making partial volume effects unavoidable. We used advanced PET reconstruction methods to reduce this issue, though residuals biases in Aβ quantification cannot be entirely excluded. Our structural analyses also benefited from surface-based analyses and advanced FW-corrected diffusion metrics, which minimize confounding from segmentation errors and non-tissue signals.^12,29,31^ In addition, our cohort lacks representation of late-stage AD individuals, which limits our ability to assess longitudinal trajectories beyond the preclinical and prodromal stage of the disease. While the focus on preclinical and prodromal stages aligns with therapeutic strategies targeting early disease mechanisms, future multi-cohort studies are needed to validate these trajectories across the entire AD continuum.

Overall, our study shows that Aβ pathology is associated with nonlinear, stage-dependent alterations in brain structure, with distinct spatiotemporal signatures across the AD continuum. By integrating data-driven regional vulnerability maps and longitudinal trajectories of brain structures alterations, we refine the interpretation of structural changes in early AD and help reconcile inconsistencies observed in previous studies.

## DATA AVAILABILITY

Data used in this study were obtained from the Pre-symptomatic Evaluation of Experimental or Novel Treatments for Alzheimer’s Disease (PREVENT-AD). Some of the data were publicly accessible at (https://openpreventad.loris.ca and https://registeredpreventad.loris.ca). Additional data can be provided upon request, pending approval by the scientific committee of the Centre for Studies on Prevention of Alzheimer’s Disease (StoP-AD) at the Douglas Mental Health University Institute.

## ACKNOWLEDGEMENTS

This work was supported by fellowship from Chinese Scholarship Council (201906070287; to TQ). The PREVENT-AD (Pre-symptomatic Evaluation of Experimental or Novel Treatments for Alzheimer’s Disease) data were funded by a Canadian Institutes of Health Research foundation grant, an Alzheimer’s Association grant, a joint Alzheimer’s Society Canada and Brain Canada Research, a National Institutes of Heath grant and a Lemaire foundation donation. The authors would like to thank Alfonso Fajardo-Valdez, Alexandre Pastor-Bernier, Bery Mohammediyan, Daniel Bowie, Jordana Remz, Mohammadali Javanray, Valentin Ourry, and Yara Yakoub for helpful discussions and feedback for this study. We are also grateful to John Breitner and Judes Poirier, founders of the PREVENT-AD cohort, who supported all MRI data acquired prior to the scanner upgrade, and to Nathan Spreng, who supported MRI data acquired after the upgrade through funding from the National Institutes of Health (R01 AG068563) and the Alzheimer’s Association (AARG-22-927100). We wish to thank all the participants and their families in the PREVENT-AD study.

## ABBREVIATIONS

AD: Alzheimer’s Disease
Aβ: β-amyloid
PET: Positron Emission Tomography
MRI: Magnetic Resonance Imaging
DTI: Diffusion tensor imaging
PREVENT-AD: PRe-symptomatic EValuation of Experimental or Novel Treatments for Alzheimer’s Disease
SUVR: Standardized Uptake Value Ratio
MCI: Mild Cognitive Impairment
FW: Free water
MD_T_: Free-water-corrected mean diffusivity

