## Supplementary Table 1 for "Tracking a decade of structural changes in preclinical and prodromal Alzheimer’s disease: insights from amyloid-β pathology"

### SUPPLEMENTARY TABLES AND FIGURES

**Table S1. MRI scanner parameters for Stage 1 and Stage 2**

| Scan type | Sequence | Stage 1 parameters (pre-upgrade) | Stage 2 parameters (post-upgrade) |
|---|---|---|---|
| T1-weighted anatomical | MPRAGE | 3D sagittal; TR = 2300ms; TE = 2.96ms; TI = 900ms; a = 9°; FOV = 256×240×176 mm; phase encode A-P; BW = 240Hz/px; GRAPPA 2; voxel size = 1×1×1; scan time = 5.12 min. | 3D sagittal; TR = 2300ms; TE = 2.98ms; TI = 900ms; a = 9°; FOV = 256×256×192 mm; phase encode A-P; BW = 240Hz/px; GRAPPA 2; voxel size = 1×1×1; scan time = 5.30 min. |
| diffusion-weighted imaging (DWI) | Pulse gradient spin echo (PGSE) EPI | 2D transversal; TR = 9300 ms; TE = 92 ms; FOV = 192×192×130 mm; phase encode A-P; BW = 1628 Hz/px. b = [0,1000] s/mm2 with 1, 64 directions; voxel size = 2×2×2; scan time = 10.15 min. | 2D axial; TR = 3000ms; TE = 66 ms; a = 90°; FOV = 220×220 mm; 81 slices; phase encode P-A; BW = 2272Hz/px; phase PF 7/8; GRAPPA 2; SMS 3; b = [0, 300, 1000, 2000] s/mm2 with [9, 7, 29 64] directions and b=0 images with phase encode A-P acquired for distortion correction; voxel size = 2×2×2; scan time = 5.49 min. |

**Figure S1. Participants included in the present study.**

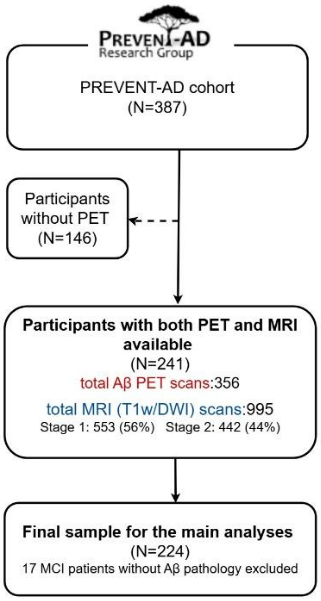


Abbreviations: PREVENT-AD, Pre-symptomatic Evaluation of Experimental or Novel Treatments for AD; dMRI, diffusion weighted MRI; MCI, mild cognitive impairment.

**Figure S2. PET/MRI acquisition dates for PREVET-AD data included in this study.**

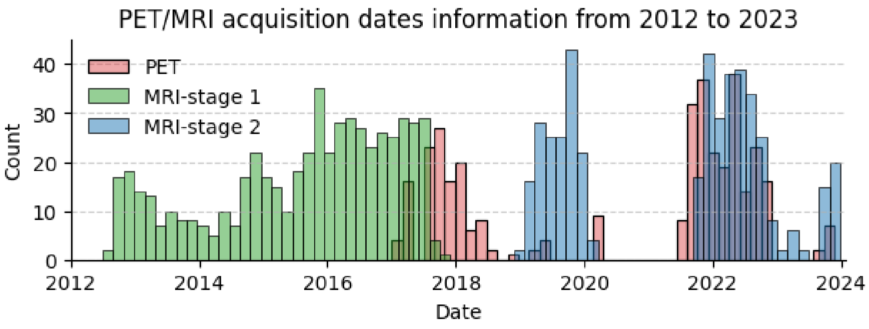


**Figure S3. Inferior parietal structural measures (brain volume, cortical thickness, cortical MD$_T$) before and after LongComBat harmonization.**

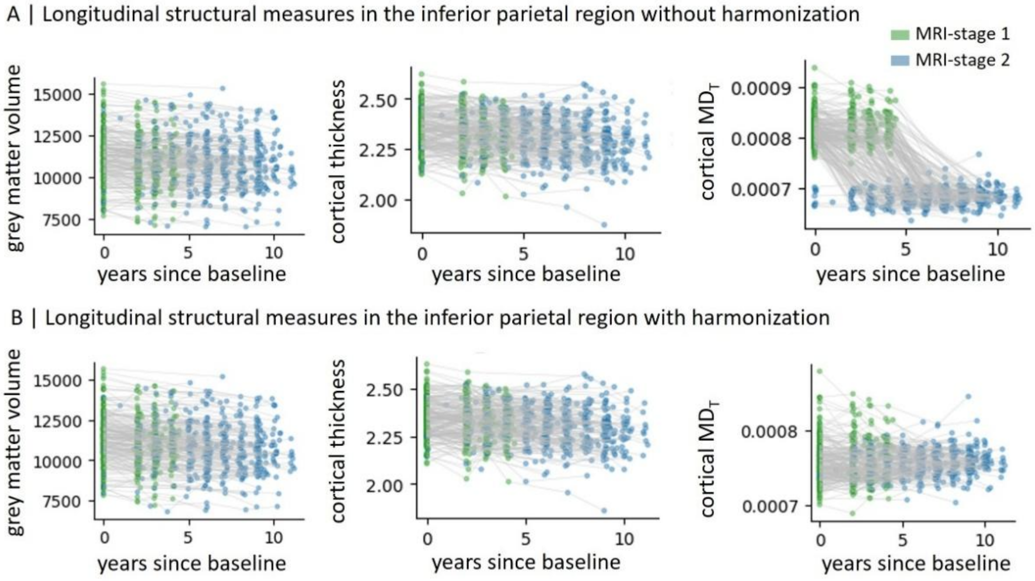


**Figure S4. PLS loadings for the second latent variable (LV2) in the Aβ− group**

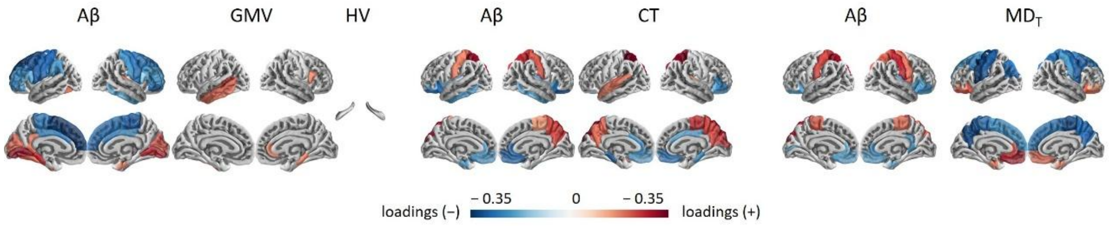


**Figure S5. PLS bootstrap ratios for the first latent variable (LV1) in both the Aβ− and Aβ+ group**

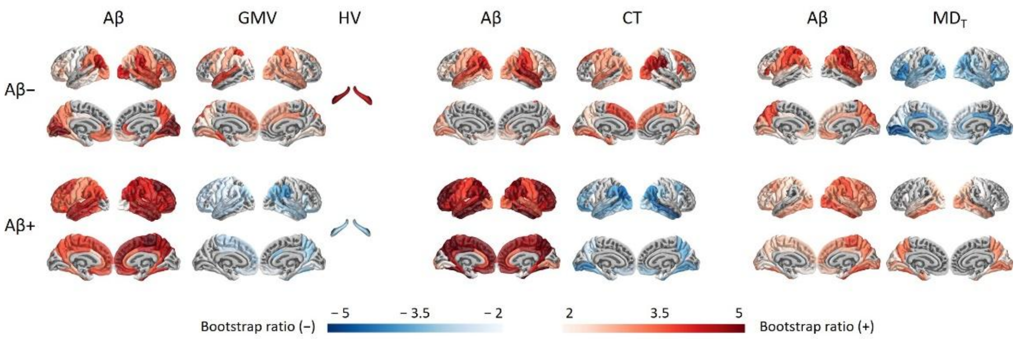


**Figure S6. Correlation between PLS-derived bootstrap ratios and loadings for LV1 in Aβ− and Aβ+ groups**

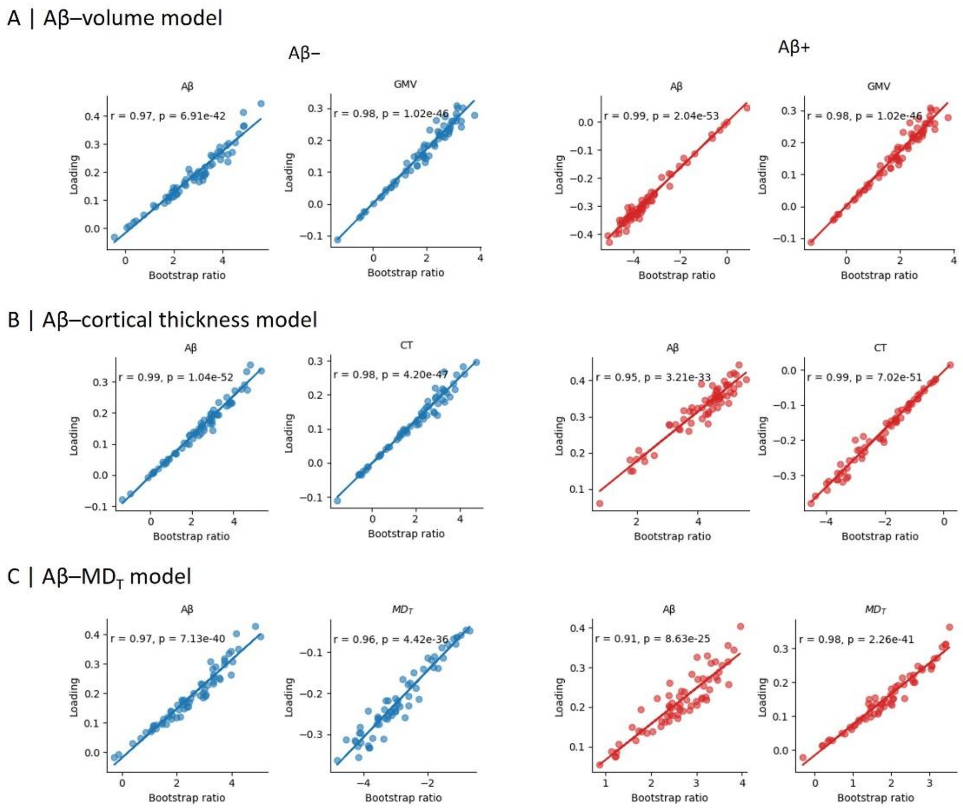


**Figure S7. Associations of Aβ rank with structural measures in cross-sectional and longitudinal PLS-derived ROIs**

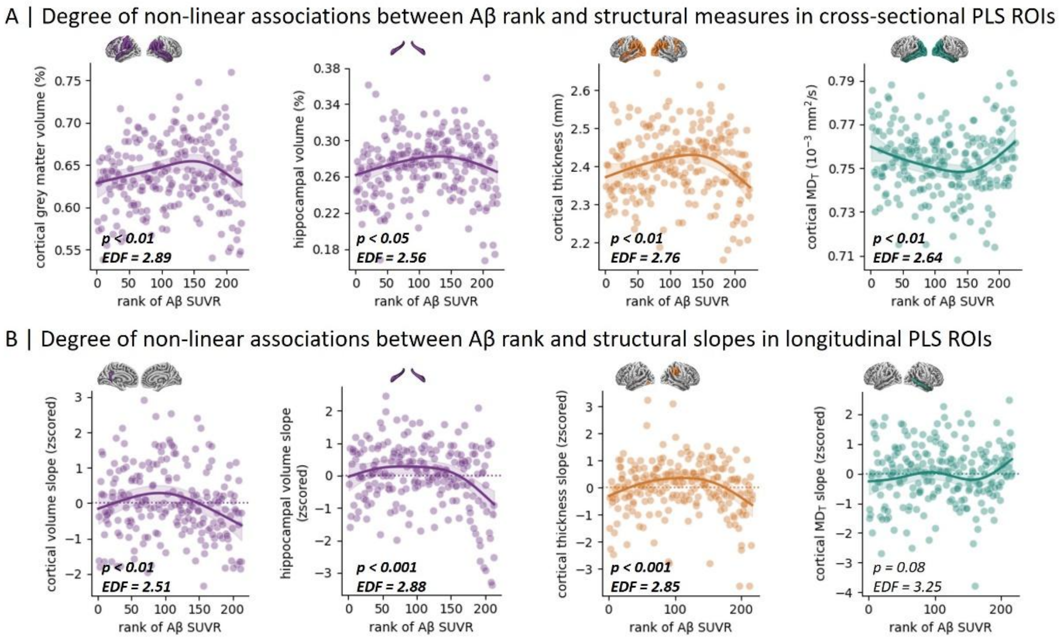


**Figure S8. Associations of Aβ and structural measures in control regions**

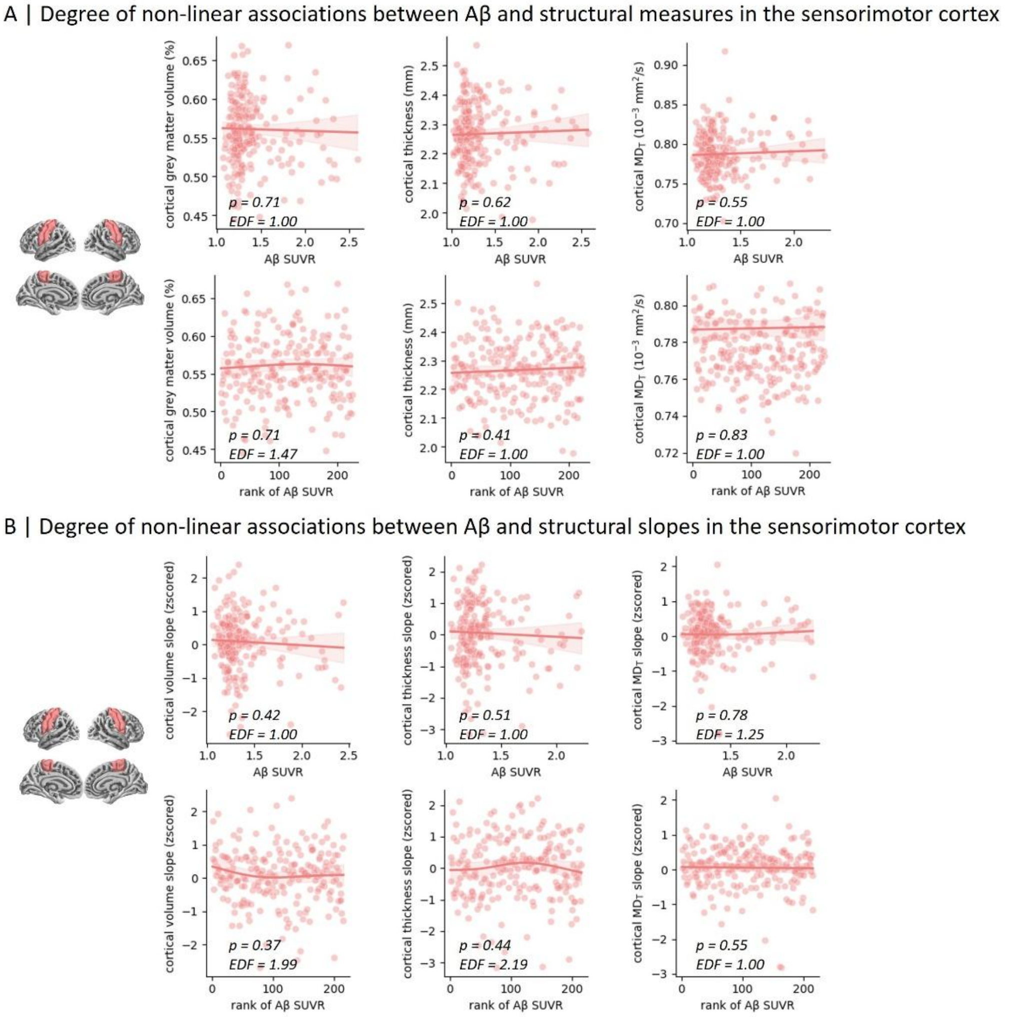

**Figure S9. Trajectory of structural changes in ROIs from cross-sectional and longitudinal PLS**

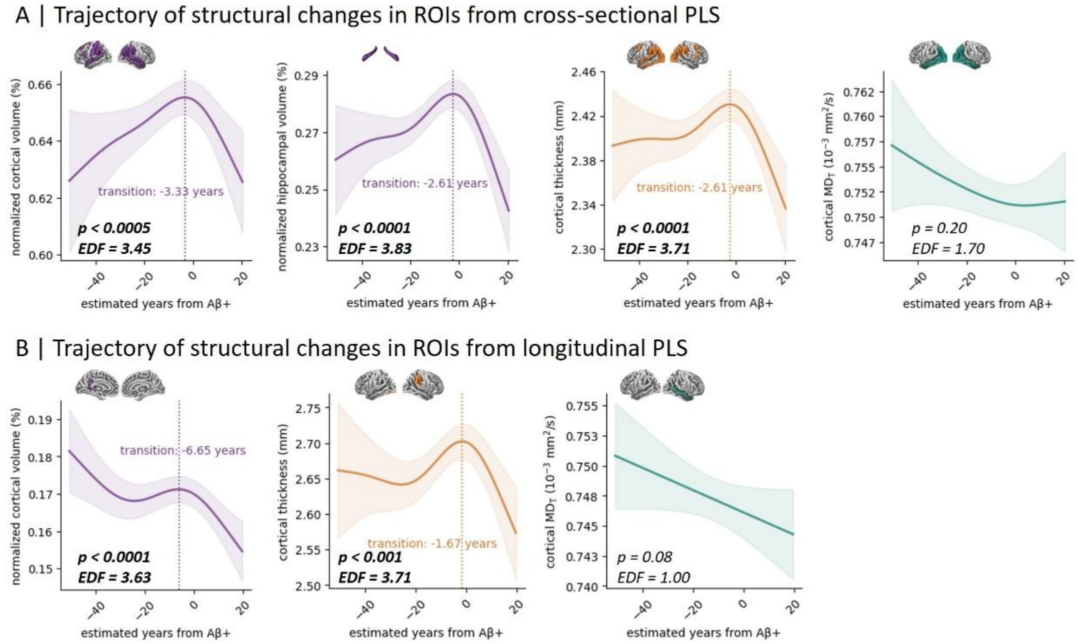


**Figure S10. Trajectory of structural changes in sensorimotor control regions**

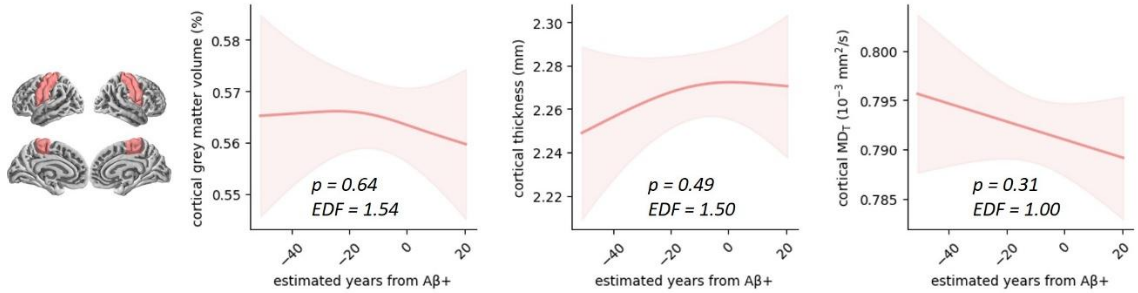
